# Characterization of longitudinal transformation of T2-hyperintensity in oligodendroglioma

**DOI:** 10.1101/2020.07.03.20145698

**Authors:** Dieter H. Heiland, Robin Ohle, Debora Cipriani, Pamela Franco, Daniel Delev, Simon P. Behriger, Elias Kellner, Gergana Petrova, Nicolas Neidert, Irina Mader, Mateo Fariña Nuñez, Horst Urbach, Roman Sankowski, Jürgen Beck, Oliver Schnell

## Abstract

**Background:** Oligodendroglioma (ODG) are CNS resistant tumors characterized by their unique molecular signature, namely a combined deletion of 1p and 19q simultaneously to an IDH1/2 mutation. These tumors have a more favorable clinical outcome compared to other gliomas and a long-time survival that ranges between 10-20 years. However, during the course of the disease, multiple recurrences occur and the optimal treatment at each stage of the disease remains unclear. Here we report a retrospective longitudinal observation study of 836 MRI examinations in 44 ODG patients.

**Methods:** We quantified the volume of T2-hyperintensity to compute growth behavior in dependence of different treatment modalities, using various computational models.

**Results:** The identified growth pattern revealed dynamic changes, which were found to be patient-specific an did not correlate with clinical parameter or therapeutic interventions. Further, we showed that, surgical resection is beneficial for overall survival regardless the WHO grad or timepoint of surgery. To improve overall survival, an extent of resection above 50% is required. Multiple resections do not generally improve overall survival, except a greater extent of resection than in previous surgeries was achieved.

**Conclusions:** Our data aids to improve the interpretation of MRI images in clinical practice.

## Background

Oligodendroglioma is the third most common type of diffusely infiltrative glioma, with an annual incidence of about 0.6 cases per 100,000 people [1] and accounts for 4-15% of all gliomas [2]. In the revised version of the WHO classification of tumors of the central nervous system of 2016, both histological and molecular parameters were included for the first time to define several glioma entities [3]. Since then, oligodendrogliomas are now characterized by a distinct molecular genotype namely the 1p19q co-deletion along with the simultaneous presence of an IDH1/2 mutation and are further graded according to their histopathological degree of malignancy [3]. Although oligodendrogliomas show highly variable clinical courses with overall survival rates ranging between 6 months and more than 20 years [4], the prognosis is relatively favorable with an average long-term survival of about 15 years. The treatment of oligodendroglioma includes surgical resection, radiotherapy and chemotherapy. Regarding first line treatment, recently, a prospective randomized trial revealed an improved overall survival for an initial sequential radio- and chemotherapy with Procabazine, Lomustine and Vincristine [5]. However, over the course of the disease, recurrences invariably occur which require further treatment planning. Against the backdrop of the relatively low incidence and the usually long course of the disease, the effects of primary applied therapy overlap with those of relapse treatment regimens and it becomes difficult to determine the particular effect of each therapy. On this basis it can be concluded that there are currently no specific therapeutic strategies with sufficient evidence for the recurrent stage of oligodendroglioma. Furthermore, the direct effects of the therapies, such as radio- and chemotherapy have only been insufficiently investigated. For example, the volume reduction of the tumor mass after surgical removal has been described by other authors [6], but the knowledge about volume reduction and growth behavior after radio- or different chemotherapies is poor.

This study aims to investigate the longitudinal growth behavior based on changes of T2-hyperintensity, to quantify the effects of different therapies on tumor growth and to identify similarities of oligodendroglioma growth patterns.

## Methods

### Study Design and Patient Cohort

We conducted a retrospective analysis of patients contained on an institutional database of WHO-grades II and III gliomas. All histological confirmed diagnosis oligodendroglioma (OG) and oligoastrocytoma (OA) were selected and re-classified in accordance to the revised WHO classification of 2016, including a molecular analysis of IDH 1/2 mutation and 1p19q co-deletion. We subsequently selected patients with a minimum follow-up of 5 years and a total number of at least 15 follow-up MRI scans. Then, a reconfirmation of the IDH mutation status and the 1p19q co-deletion was performed, resulting in 44 patients who were finally enrolled in the study. All patients received a neurosurgical intervention at the Department of Neurosurgery, University Hospital Freiburg, Germany between 2000 to 2018. An informed consent for the scientific exploration of clinical and biological data consistent with the local ethical standards and the Declaration of Helsinki was available from all patients. The study was approved by the ethic committee University of Freiburg (protocol 472/15_160880). The methods were carried out in accordance with the approved guidelines.

### Histopathological and Molecular Diagnostics

Tissue samples were fixed using 4% phosphate buffered formaldehyde and paraffin-embedded according to standard procedures. Analysis were performed in the Institute of Neuropathology, Medical-Center University of Freiburg as detailed described in our previous works[7, 16].

### Tumor Segmentation

Tumor growth was analyzed by tumor segmentation performed using the “NORA” software tool, a web-based framework for medical image analysis developed by the Department of Radiology, Medical Physics, University of Freiburg (http://www.nora-imaging.com). T1-weighted images with contrast-enhancement as well as T2- and T2-FLAIR-weighted images were analyzed. Since part of the patients, in particular in the early follow-up years (from 2000 to 2008), had not received 3D datasets, a reconstruction of the given axial, coronal and sagittal images was necessary to achieve the best possible segmentation. We performed tumor segmentation of contrast-enhanced regions if existing, as well as FLAIR hyperintense regions. All segmentations were performed by trained specialists in an *in-silico*-assisted manual manner, which means that the tumor areas were supervised, and the algorithm calculated the boundaries according to the supervised markings. The temporal course of the disease regarding tumor resection and treatment modalities was reconstructed by accessing clinical documentation and aligned to the time-points of follow-up images resulting in a table containing all clinical and image information (**Supplementary table 1**.).

### Normalization and Fitting

We started the model by normalization of all patients into a time-dependent multidimensional matrix containing tumor volume in the T2-weighted as well as in the T1-weighted contrast enhancing images, time point of surgical procedures and time interval for radio- and different chemotherapies. We centered an z-scored (1) each individual volumina and fitted the growth curves by loess-fit (2) from the stats-package (R-software) to extrapolate the gaps between follow-up timepoints.

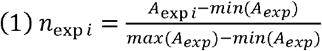

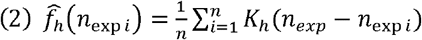

K is the kernel and 0.8 > h > 0.2 is used to adjust the estimator. The model is used for further evaluation of therapy modalities.

### Model Extent of Resection

The extent of resection was defined based on the presurgical volume: MRI_(ts-1)_ (ts is timepoint of surgery) and the postsurgical MRI within 3 months: *mean*(MRI_(ts+1…n)_) (n contained all MRI within 3 months). In some cases, direct postoperative images or early postoperative MRI showed more frequently unspecific T2-hyperintensity artefacts. In order to overcome this bias, we used the mean of all MRIs within 3 months postoperative. Differences in extent of resection was tested by Mann-Whitney-U-Test, significant was determined by p<0.05.

### Model of Therapy Responses

Therapy response was computed by the dynamic changes during a period of 1.5 year after initial treatment. We used the loess-fit model described above to compare dynamic changes as response of a treatment. A “Partitioning Around Medoids” (PAM) cluster was used to group all responses based on their similarity. We used gap-statistics to compute the optimal number of clusters.

### Cox-Regression Model

Analysis were performed using the “survival” package in R-software in order to estimate overall survival and log-rank tests to compare our cohort or cluster groups. Hazard ratios and 95% confidence intervals were estimated performing a Cox proportional-hazards regression model including a 10-fold cross-validation. We determine the alpha-level at 5% to achieve statistical significance with a power of >80%. Patients who continue to live or whose survival is not evident are censored in the analysis, a detailed description is given in a recent publication[17].

### Cox-Regression Model with Time-Dependent Covariates

As detailed described in our recent work, we were challenging the meaningfulness of a classical Cox proportional-hazards regression model. The classical model is strongly biased due to the fact that only patients with improved survival may receive an additional therapy at a later stage and most of the patients were censored but hat multiple recurrences. To overcome the limitations of a classical Cox regression model, we included all recurrences and added a time-depended-covariates to the model. By splitting the variable ‘recurrences’ into its individual time intervals we were able to individually analyze therapeutic responses within the duration of the whole course of the disease. Additionally, multivariate regression was performed including outcome-dependent variables, a detailed description is given in a recent publication[17].

## Results

### Patient Cohort and Molecular Reclassification

We started our investigation by screening the Medical-Database Freiburg, resulting in 212 patients who matched the diagnosis of oligodendroglioma (OG) and oligoastrocytoma (OA), from which 96 were correctly re-classified as true OG according to the 2016 revised WHO classification of tumors of the central nervous system. In order to warrant consistent data quality, we sorted out all patients who were not able to reach the quality criteria we defined (methods part). For further analysis, we enrolled 44 patients, which reflected the low incidence of molecular defined Oligodendroglioma. Most frequently the tumor was observed in the frontal lobe (n=35), here also localizations with multi-lobular extension were counted. Less often were temporal (n=11), parietal (n=8) or occipital (n=2) localizations, **Figure 1a**. In our cohort, gender and age was well balanced similar to previous works[7].

**Figure 1:**
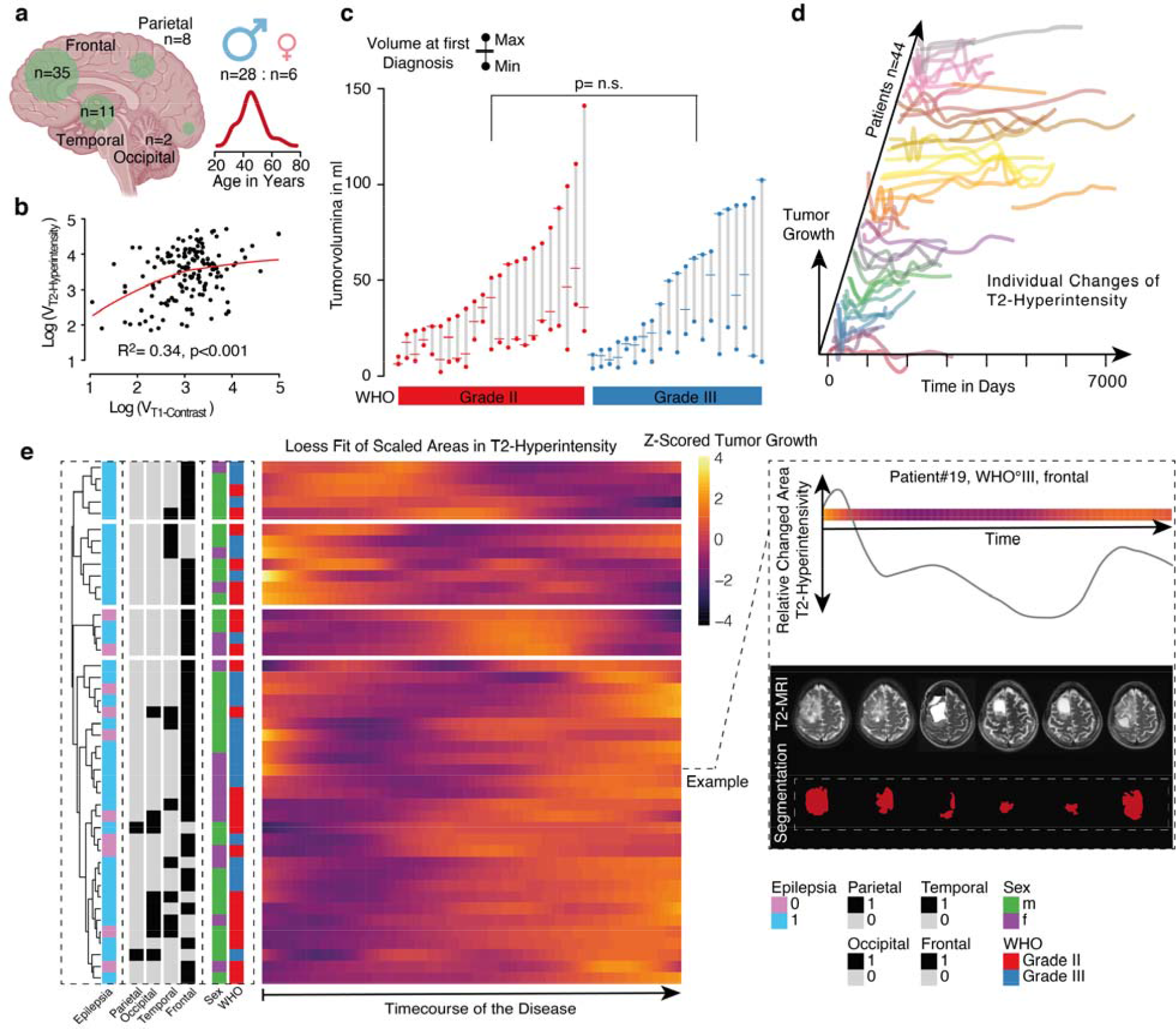
**a)** Overview of major characteristics of the cohort, distribution of age, gender and tumor localization **b)** A scatter plot indicate the correlation between T1 contrast enriched volume and T2-hyperintensity volume. **c)** A waterfall plot illustrates the minimal and maximal volume (T2-hyperintensity) and the volume at first diagnosis, colors indicate the determined WHO grade. **d)** Line plots show individual growth curves based on T2-hyperintensity changes over the course of the disease. Each patient is colored differently. **e)** Heatmap of loess fitted growth curved (Time∼T2-hyperintensity changes), darker colors show less growth, brighter colors indicate timepoints of increased growing. At the left side, clinical characteristics are aligned to each patient (as rows). On the right side, an example of one patient was shown in order to improve understanding of the heatmap, including the heatmap, growth curve and corresponding T2-MRI images. At the right bottom, the color code for clinical parameters is given.

### T2-Hyperintensitivity Reflect Longitudinal Tumor Growth

In order to investigate longitudinal changes of tumor growth we aimed to identify an MRI surrogate parameter, which on the one hand should be widely available and on the other hand be contained in MRI recordings at the early stages of some long-term survivors (year ∼2000). Due to the large heterogeneity of available images with regard to image quality, various MR scanners and multiple MR-protocols, only a limited number of MR sequences could be considered for further analyses.

We measured the T2-hyperintensity, FLAIR-hyperintensity, T1 +/− contrast and found an expectable correlation between FLAIR and T2 hyperintensity whereby, the T2-hyperintensity was measurable within 96.2% of all imaging records while FLAIR imaging was in less than 80% available (no sagittal FLAIR sequences). Although the T2 hyperintensity does not allow for a doubtless determination of the tumor volume, the error within the entire cohort is relativized. Our data contained an expectable low frequency of T1-contrastenrichment of the ODG (34%) which clearly indicate active tumor regions (pre-treatment), in contrast, changes of the T2 hyperintensity are susceptible to non-specific changes and not indicate active parts of the tumor. When comparing volume from T1 contrast-enrichment and T2 hyperintensity, a relatively high correlation was obtained (R^2^=0.34, p<0.001), **Figure 1b**. We assume that based on our available data, T2 hyperintensity volume can be sufficiently used to determine the volume of OGD in a longitudinal observation study.

### Diversity of tumor size and longitudinal volume transformation

In a next step, we mapped minimal and maximal tumor volume of our patients and found a strong variance ranged between 10.16ml and 141.06ml (maximal tumor size), **Figure 1c**. No significant difference was found between WHO grad II and III patients (WHO grade II: 54.13ml vs. WHO grade III: 51.65ml p=0.79). By comparison of the individual growth curves, determined by the longitudinal changes of T2-hyperintensity volume, we observed a pronounced heterogeneity of growth behavior, **Figure 1d**. In order to find similarities in the behavior, we performed clustering, which resulted in 3 different patterns, **Figure 1e**. As expected, the growth peak (measured as maximum volume) can be found either at the beginning, during or at the end of the disease. We were not able to identify any significant accumulation of clinical parameters in these clusters, nor was there any difference between WHO grade II or III tumors. Hence, we suspect that these differences are due to various response to the received treatments and intend to investigate this in greater detail in the following sections.

### Extent of resection and surgical treatment

The surgical treatment of OGD is undisputed and associated with a significant increase in overall survival. However, it remains unclear when is the best time for resection and how effective is resection at the recurrent stage? Is prior chemotherapy or radiotherapy beneficial?

In order to address these questions, we analyzed 61 resections from 38 patients (6 patients underwent only biopsy before adjuvant treatment). The number of surgeries per patient ranged between a single to 4 resections over the course of the disease. We observed a median reduction of the T2-hyperintensity volume from 57.8ml to 24.4ml (57.78%, p=0.0018) and a reduction of T1-contrast enriched volume from 3.54 ml to 0.564ml (84.06%, p=0.0051), **Figure 2a-b**. Four patients showed a pronounced volume after resection due to unspecific T2-hyperintensity increase in early postoperative imaging. From a surgical point of view, not all tumor locations are equally accessible, some localizations are more approachable than others. In our cohort 64.4% of the resections were performed at the frontal lobe which is relatively easily accessible. In comparison to other localizations, however, there is no improved extent of resection (measured in volume reduction) compared to frontal tumors (reduction frontal 58.23% vs. non-frontal 52.5%, p=n.s.), **Figure 2c**. In our cohort, only 17.85% of the patients (n=10) received a resection at first diagnosis, **Figure 2d**, the extent of resection did not differ between initial resection or resection at a later time point with previously received treatments (p=n.s.), **Figure 2e**. Further, we observed an expected significant difference between primary and recurrent resections (p=0.013) with lower extent of resection in the recurrent stage of the disease, **Figure 2f**. No difference regarding the extent of resection was observed between WHO grad II and III ODG (p=n.s.), **Figure 2g**. In summary, resection can be performed without significant differences of extent of resection at multiple timepoints during the disease, the extent of resection measured by T2 hyperintensity is lower than reported for malignant tumors, which is expectable due to the infiltrative nature of the disease. A conclusion on how patients benefit from resections at varying timepoints is not possible without considering the other therapy modalities and will be examined in the following in a multivariate analysis.

**Figure 2:**
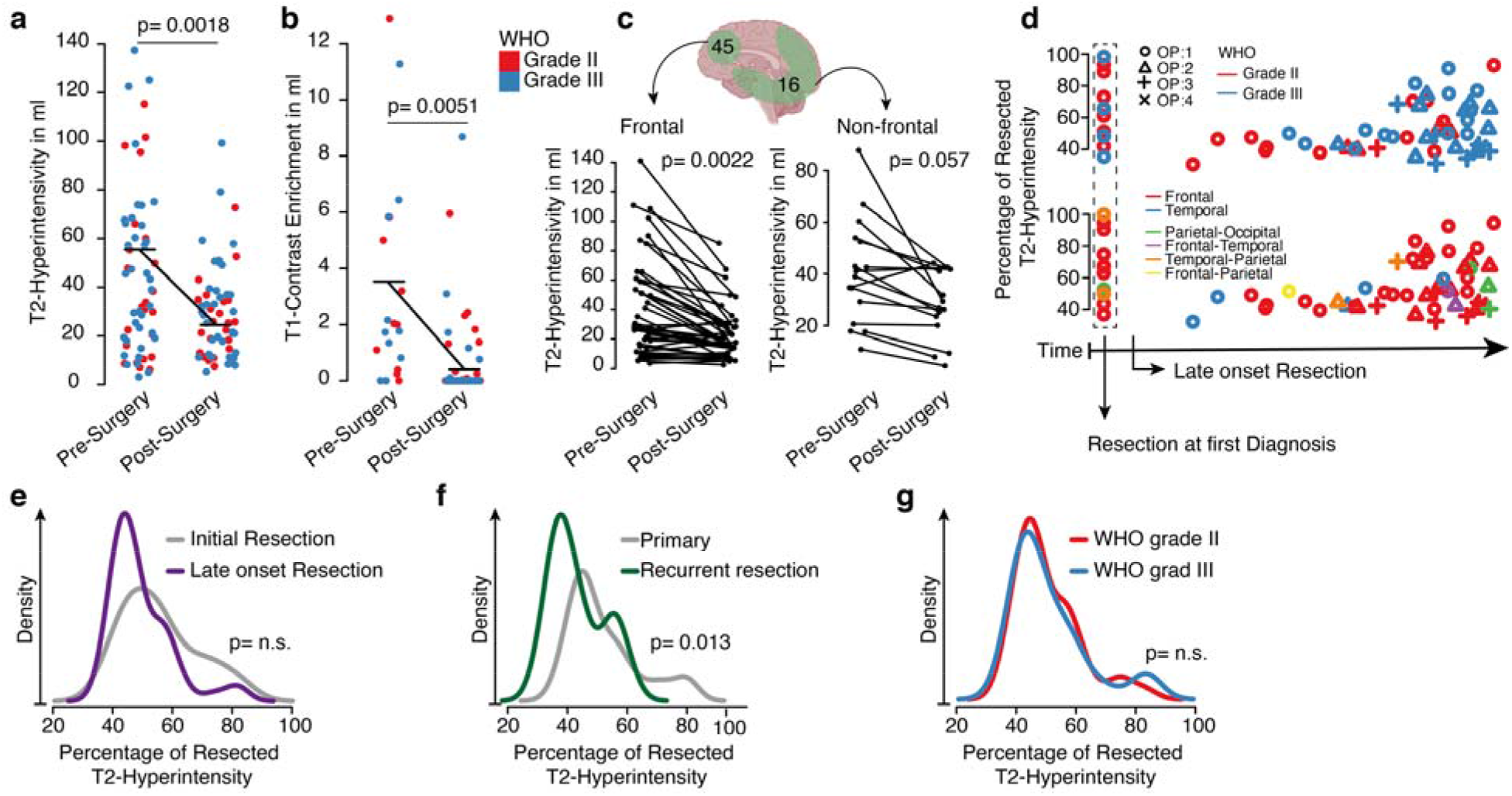
**a-b)** Scatter plot of T2-hyperintensity volume (a) and T1-contrast enrichment volume (b) pre- and postsurgery of ODG. Colors indicate the WHO grade and black lines illustrate median difference between pre- and postsurgery. significance was tested by Wilcoxon Rank Sum test. **c)** Connected lines illustrate the difference between pre- and postsurgery volume in frontal (n=45) surgeries and non-frontal (n=16) surgeries, significance was tested by Wilcoxon Rank Sum test. **d)** Scatter plot of time-dependent (x-axis) differences of extent of resection (y-axis percentage of resected tumor). Shapes indicate the number of surgeries and colors indicate WHO grade (upper plot) or localizations (bottom plot). **e-f)** Density of percentage of resection (x-axis) between different groups as indicated by the colors.

### Adjuvant and Neo-adjuvant Treatment of OGD

In addition to surgical treatment, radio- and chemotherapy represent the second pillar of OGD therapy. The large variance of chemotherapies in use substantially complicates the accurate evaluation of individual effects of chemotherapy. So far, the standard chemotherapeutic agent is a combination of Procabazine, Lomustine (CCNU) and Vincristine (PCV), due to toxic effects, especially neurotoxic effects of vincristine, often only PC (Procabazine, Lomustine) was used. Temozolomide (TMZ) has also been widely used for therapy, especially in later stages of therapy and in recurrent tumors. In order to obtain an overview of the therapeutic effects, we recorded the T2-hyperintensity dynamic from the initiation of chemotherapy (79 adjuvant treatments of 44 patients) over 2 years and sorted into 3 groups by clustering, **Figure 3a**. In order to obtain a more accurate impression of the T2 changes within the clusters, we have traced the mean growth-curves indicating that cluster 1 (n=47 treatments, 59%) demonstrates a constant growth in T2-hyperintensity volume after therapy (PC 34%, Radiotherapy (RT), 23%, TMZ+RT 23%, TMZ 14%, PCV 2%). However, in addition to this most anticipated growth behavior, two additional clusters appeared, which reflect the complexity of MRI interpretation in the treatment of ODG. Approximately 40% of the administrated therapies do not fit into the expected response and showed an early “pseudo”-progress (early T2-hyperintensity increase) followed by an agitated high dynamic response (generally lower volume compared to initial, pre-therapeutic volume, cluster 2) or a likely linear decrease of volume (cluster 3), **Figure 3b**. Across all clusters, no significant enrichment of clusters or clinical features was obtained, **Figure 3c**. In summary, here we described the different responses to adjuvant- and neoadjuvant treatment, showed that volume changes are highly dynamic and only approximately half of the patient showed the expected response pattern.

**Figure 3:**
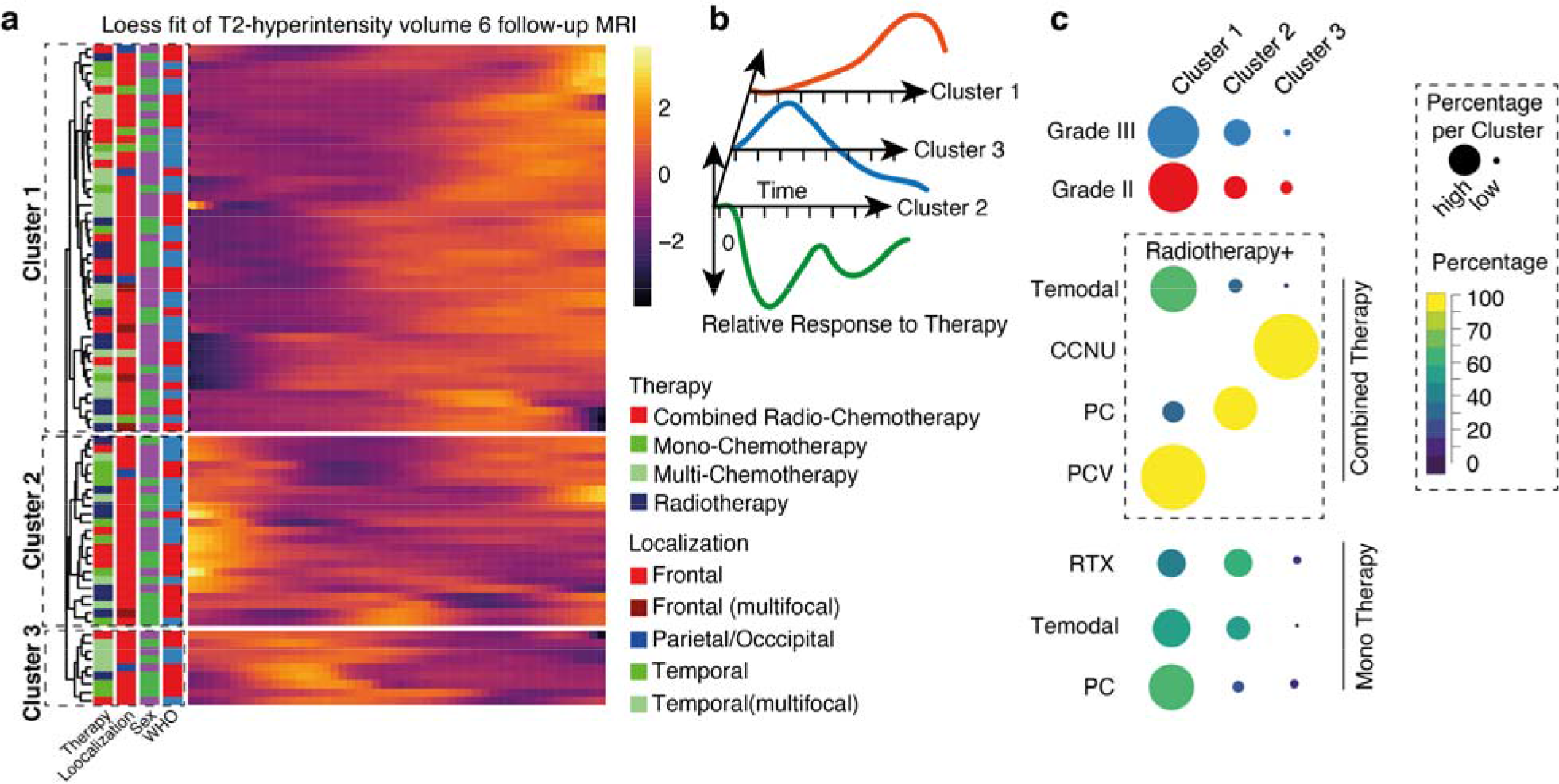
**a)** Heatmap of loess fitted growth curve of a two years interval after initiation of radio- or chemotherapies (Time∼T2-hyperintensity changes), darker colors show less growth, brighter colors indicate timepoints of increased growing. At the left side, clinical characteristics are aligned to each therapy (as rows). At the right bottom, the color code for clinical parameters is given. **b)** Mean fitted curve of patients with similar growth behavior merged into cluster 1-3. **c)** Percentage of therapies or clinical features in each cluster, illustrated as dots (colored according to percentage) with size according to percentage, larger dots indicate higher percentage.

### Adjuvant and Neo-adjuvant Treatment of OGD

Based on our gained information regarding the heterogenous response in general growth behavior and to different treatment modalities, we finally aimed to identify parameter that have impact on the course of the disease. We used a multivariate Cox proportional-hazards regression model with tumor relaps as a time-depended-covariates, which allowed us a more precise prediction of survival relevant parameters. First, we analyzed to what extent different growth properties as described in the first part of the analysis impact overall survival. We were not able to identify a growth pattern which was associated with beneficial clinical outcome, **Figure 4a**. Additionally, WHO grade, numbers of resection (more or less than 2), gender and observed seizures during the disease did not change outcome of the patients, **Figure 4 b-d**. Next, we validated the extent of resection and found a significant improvement of survival in patients with less residual tumor then 25% and 10%, **Figure 4e**. All other parameters (including chemo- or radiotherapy) did not reach significance. Further, we used the overall survival and the extent of resection (maximal extent in patients that receive multiple resections) to create a prediction model. By use of this model, we demonstrated that increased overall survival was obtained in patients with less than 50% of residual tumor.

**Figure 4:**
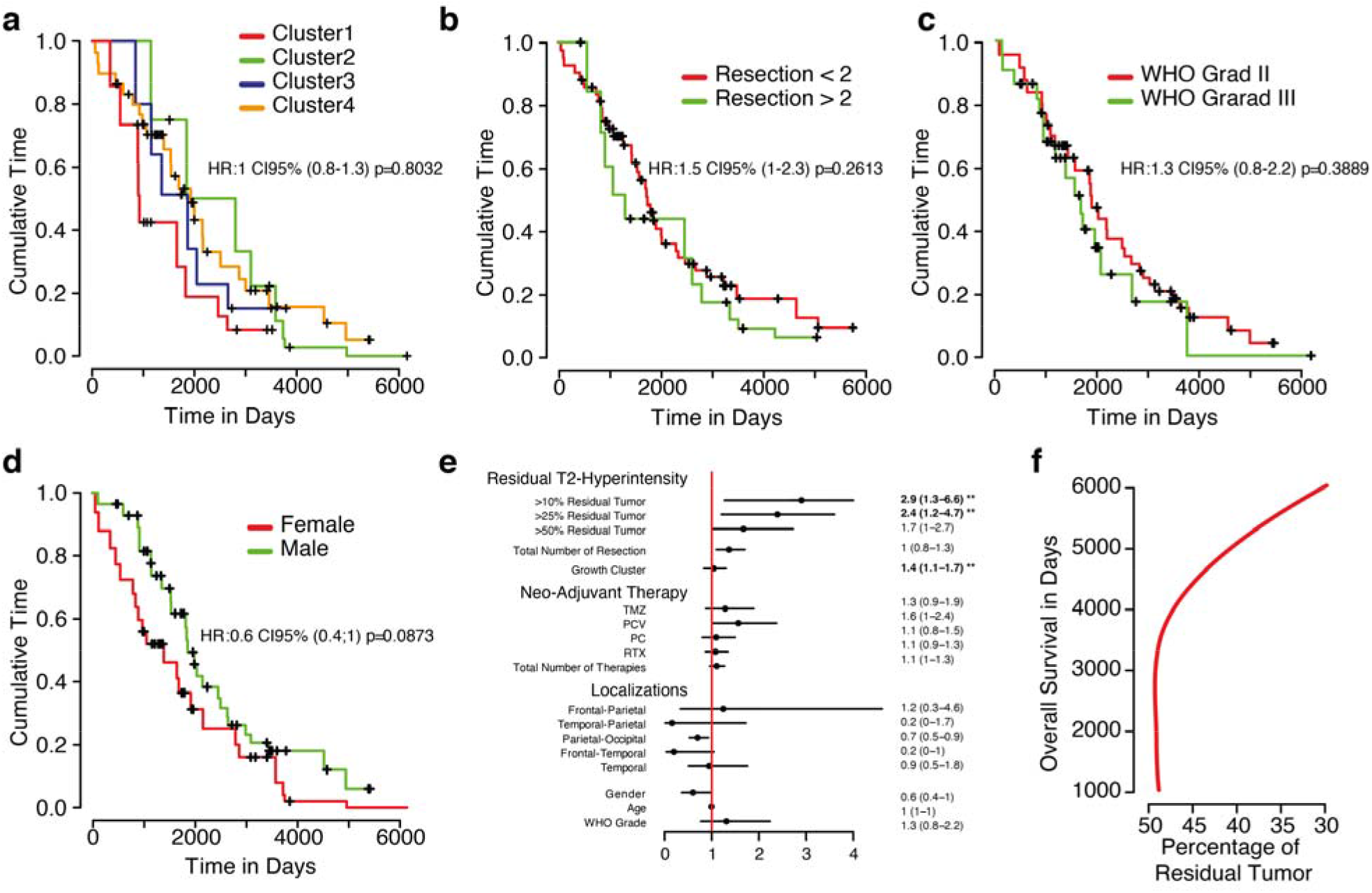
**a-d)** Survival curves based on Kaplan-Meier statistics for growth pattern of T2-Hyperintensity (a) number of resections (b), WHO grade (c) and Gender (d). **e)** Cox-Regression of multiple clinical features, *p<0.05, **p<0.01, **f)** Loess model of overall survival ∼percentage of residual tumor.

## Discussion

The proper oncological treatment of oligodendroglioma represents a challenge due to the highly variable clinical course of the disease with survival rates between few years and more than two decades [4]. Two crucial prospective trials over the past few years showed a favorable response to combined radio- and chemotherapy in WHO grade III oligodendroglioma and oligoastrocytoma [8, 9]. One drawback of these important studies is the lack of detailed molecular information, which hampers the result interpretation into the spotlight of the revised version of the WHO classification. Furthermore, there is still an ongoing debate to which extent WHO grade II tumors should be treated, especially in young patients. In this case, the maintenance of quality of life should be one of the highest priorities and needs to be weighed against the risks and benefits of treatment [10]. Here we report a relatively small cohort of 44 molecular defined oligodendroglioma, reflecting the rarity of this tumor entity. Although the limited number of patients in our analysis, we were able to achieve a total number of 836 MRI images which allowed us to map the transformation of tumor volume over time. Based on these findings we observed a strong variability regarding time-depended changes of tumor volume without significant association to clinical features. This reflects daily practice, whereas dynamic changes of tumor volume difficult to interpretative and severely hamper clinical decisions. We identified that this dynamic pattern could be also observed after different therapies without correlations to a specific therapy.

Although recent trials investigated specific treatment regimens, such as initial sequential radio- and chemotherapy with Procabazine, Lomustine and Vincristine [5] or the role of surgery [6, 7], the impact of those therapies in the recurrent stages of the disease remains questionable. In the recurrent stage of advanced disease, there is a lack of evidence of which therapies provide the greatest benefit, and there are no valid instruments for monitoring the success of therapy.

We were not able to identify different responses of chemo- or radiotherapy when administrated initially or during the disease. However, we did not observe a benefit of a single chemo reagent likely due to multiple therapies which strongly bias the effect of a single treatment. Our cohort, each patient was at least treated with two treatment modalities, therefore the evaluations regarding individual therapies could not to be used.

Surgery of oligodendroglioma are also controversy discussed, especially in the later stage of the disease and if only a small amount of the tumor is respectable due to eloquent localization. Recent studies showed a clear significant improvement of overall survival in resected patients[11–13], but recurrent surgeries, however, multiple resections in OGD revealed no additional benefit for the patient[7]. Our work can largely confirm these obtained results. Further, in contrast to different chemotherapy regimens, resection is able to stand out with significant survival improvement although the data are relatively fuzzy and biased through multiple therapies. In contrast to published data[7, 14, 15], we found that patients benefit from a resection if the residual tumor is less than 50%. Based on our results we would like to emphasize that the success of a resection is not binary but rather a function that reflects a negative correlation between survival and residual tumor. From this perspective it seems to be more important to perform safe resections to ensure maximum preservation of neurological function, but if less than 50% can be resected the patient does not seem to have any benefit.

The study contains numerous limitations starting with the small number of patients due to the overall low incidence of oligodendrogliomas and the monocentric character of the study. To better characterize the results, molecular data would be essential to explain the underlying mechanisms of the different growth behavior. Another bias are different surgical treatments of temporal, frontal or parietal localized tumors. In general, we were not able to find a relationship between the extent of resection and the overall survival of the patients due to multiple resections in the course of the disease. On the basis of our findings, we plan to further investigate the molecular architecture that could explain the difference of temporal and frontal oligodendrogliomas. A general limitation of our and other studies comparing novel established classifiers against the WHO classification in a retrospective setting. Due to the fact that patients received grade-dependent therapy, a correct comparison is impossible. Nevertheless, there aren’t any more accurate methods available to provide an unbiased comparison in these rare tumors.de dependent therapy makes a correct comparison impossible.

## Conclusions

We conclude with two major findings:

1. OGD are rare tumors which demonstrate a highly dynamic growth pattern with variable changes of T2 MRI imaging. Changes are individual and do not correlate with clinical parameter of therapeutic interventions.
2. Surgical resection is beneficial for overall survival, but the time point of resection (initial vs. late onset) is not important. For each resection, “the more the better”, but at least 50% of the tumor needs be resected to achieve an improvement for the patient. Multiple resections do not improve the patient survival, except it is possible to achieve a greater extent of resection than in previous surgeries.

## Data Availability

no available in preprint version

## ABBREVIATIONS

GBM: Glioblastoma
ODG: Oligodendroglioma
OA: Oligoastrozytoma
CNS: Central Nervous System
WHO: World Health Organization
TMZ: Temozolomide
PC: Procabacine + Lomustine
PCV: Procabacine + Lomustine + Vincristin

## DECLARATIONS

### Ethics approval and consent to participate

An informed consent for the scientific exploration of clinical and biological data consistent with the local ethical standards and the Declaration of Helsinki was available from all patients. The study was approved by the ethic committee University of Freiburg (protocol 472/15_160880). The methods were carried out in accordance with the approved guidelines.

### Consent for publication

Not applicable

### Availability of data and material

The datasets used and analyzed during the current study are available from the corresponding author on reasonable request.

### Authors’ contributions

All authors have read and approved the manuscript. Collection of data, performing of tumor volumetry, writing of portion of the manuscript: RO; writing of portion of the manuscript: DC, PF; conception and study design, analysis and interpretation of data, writing of portion of the manuscript: DHH; submitting of manuscript: PF; development of the software for tumor volumetry: EK; providing of imaging, interpretation of imaging, contribution to editing of the manuscript: IM, HU; analysis of neuropathology: R.S.; contribution to editing of the manuscript: O.S., D.D., S.P.B, G.P., N.N., J.W., M.T.F.-N., J.B.;

## Acknowledgements

No

## Figure legends

**Supplementary Table 1:** Clinical baseline data of the cohort.

